# Wastewater-Based Genomic Surveillance of SARS-CoV-2 Variant Circulation in Two Informal Urban Settlements in Nairobi, Kenya

**DOI:** 10.64898/2026.03.23.26349096

**Authors:** Leonard Kingwara, Rukia Sarah Madada, Vera Morangi, Shaline Akasa, Victor Kipruto, Jully Okonji, Muthoka Richard, Charles Rombo, Kanana Kimonye, Emmanuel Okunga, Moses Masika, Edwin Ochieng, Rufus Nyaga, Osborn Otieno, Fatim Cham, Noah Hull, Kamene Kimenye

**Author notes:** Co-First authors (Leonard Kingwara, Rukia Sarah Madada).

## Abstract

**Background:** SARS-CoV-2 genomic surveillance data remain limited in most low and middle-income countries (LMICs), resulting in significant gaps in the understanding of variant circulation and evolution. Wastewater-based epidemiology (WBE) presents a non-invasive, cost-effective, and population-representative surveillance approach that can complement clinical testing, particularly in densely populated urban informal settlements with limited healthcare access. This study aimed to pilot wastewater-based genomic surveillance as a multifaceted public health tool in Kenya.

**Methods:** A prospective study was conducted using wastewater samples collected from two WHO-validated environmental surveillance sites — Eastleigh A (Kamukunji sub-county) and Mathare (Starehe sub-county) — in Nairobi, Kenya, between December 2022 and October 2023. A total of 272 samples were collected using Moore swabs at a frequency of two to three times per week. Samples were concentrated using Nanotrap® Magnetic Virus Particles, and nucleic acid was extracted using the Qiagen QIAamp Viral RNA Mini Kit. SARS-CoV-2 was detected using RT-PCR (TaqPath™ COVID-19 CE-IVD RT-PCR Kit). Library preparation for whole-genome sequencing was performed using the Illumina COVIDSeq kit, and sequencing was conducted on the Illumina MiSeq platform. Bioinformatic analysis was performed using Terra.bio and RStudio, and phylogenetic analysis included sequences abstracted from GISAID.

**Results:** Of 272 samples, 238 (87.5%) tested positive with a cycle threshold (Ct) value of less than 36. Genomic analysis of 181 sequences identified Omicron as the predominant circulating variant, detected in 59% of samples. Other variants included XBB (16%), XBB.2.3(10%), XBB.1.9.X (5%), and additional minor variants. These findings were concordant with clinical sequencing data from Kenya over the same period.

**Conclusions:** Wastewater-based genomic surveillance reliably reflected SARS-CoV-2 variant trends observed in clinical data. This approach provides early signals of variant emergence and evolution, offering a cost-effective complement to clinical surveillance in resource-limited settings.

## Introduction

The coronavirus disease 2019 (COVID-19), caused by severe acute respiratory syndrome coronavirus-2 (SARS-CoV-2), remains a global public health challenge. As of March 2024, more than 700 million confirmed cases and over 7 million deaths had been reported worldwide (1). COVID-19 was first reported in Wuhan, China in late 2019 and was declared a public health emergency of international concern in early 2020 (1). Kenya reported its first case in March 2020 and had documented over 340,000 confirmed cases by April 2024 (2). Since its emergence, SARS-CoV-2 has mutated extensively, generating new variants with altered transmissibility, immune evasion potential, and disease severity (3).

According to the Global Initiative on Sharing All Influenza Data (GISAID), the first SARS-CoV-2 variants identified in Africa were B.1 and B.1.1.7, both detected in January 2020 (4). Circulating variants have evolved progressively since then, with the Delta variant (B.1.617.2) being the longest-circulating variant, persisting for over two years, and the BA.1.1 subvariant having the shortest circulation duration of approximately four months (4). In Kenya, several Delta variants and Omicron subvariants were reported to be in circulation between 2021 and early 2024 (4).

Genomic surveillance is critical for monitoring viral evolution, characterizing variants of concern (VOCs), and informing public health response strategies (5). However, conventional clinical surveillance has inherent limitations in settings with constrained healthcare infrastructure, particularly in densely populated urban informal settlements. Nairobi accounted for over 30% of Kenya’s confirmed COVID-19 cases (2), a figure that likely represents an underestimate due to limited healthcare access and the inability to capture asymptomatic infections through routine clinical testing (2).

Wastewater-based epidemiology (WBE) has emerged as a validated and scalable complementary surveillance approach. Viral RNA shedding in feces occurs in both symptomatic and asymptomatic individuals, often preceding the onset of clinical symptoms, thereby enabling detection of SARS-CoV-2 in wastewater days before clinical cases are reported (6)(7). WBE overcomes the underdiagnosis and underreporting associated with clinical surveillance and has provided valuable insights into the genetic diversity and emergence of variants of concern (8) (9). This approach is particularly relevant in informal urban settlements where clinical monitoring is logistically challenging (9).

This study aimed to address existing knowledge gaps in SARS-CoV-2 diversity in Kenya by tracking SARS-CoV-2 variant evolution in wastewater samples collected from two densely populated informal settlements in Nairobi — Eastleigh A (Kamukunji sub-county) and Mathare (Starehe sub-county). The specific objectives were to: (i) detect and characterize SARS-CoV-2 variants in wastewater over time; (ii) compare variant profiles between the two surveillance sites; (iii) correlate wastewater findings with clinical sequencing data from Kenya; and (iv) demonstrate the utility of WBE as a complement to clinical surveillance in resource-limited settings.

## Methods

### Study Design and Setting

This was a prospective observational study conducted at the National Genomics and Molecular Surveillance Laboratory (NGMSL), Kenya, between December 2022 and October 2023. Wastewater samples were collected from two WHO-validated environmental surveillance sites in Nairobi: Eastleigh A (Kamukunji sub-county) and Mathare (Starehe sub-county). These sites were purposively selected based on population density, sewer system coverage, number of schools and health facilities, settlement type, and historical COVID-19 case burden.

#### Kamukunji (Eastleigh A)

This sub-county is divided into five administrative wards: Airbase, Eastleigh South, Eastleigh North, California, and Pumwani. It is among the most prominent commercial hubs in Kenya and includes some of the oldest residential areas (Shauri Moyo, Bahati, and Eastleigh). The Eastleigh A wastewater catchment area falls within the Airbase ward, which covers an area of 9.8 km^2^, houses five health facilities, and had a projected population of 65,703 (34,199 male; 31,504 female) comprising 66,756 households in 2022. The sample collection point is located at the convergence of the main sewer line of the Nairobi Water and Sewerage Company, situated between 9th, 10th, 11th, and 12th Streets on the 2nd and 3rd Avenues (coordinates: Latitude −1.276012°, Longitude 36.852754°), covering sewage from residential, commercial, and healthcare areas from 1st to 10th Street. This site has consistently met the WHO enterovirus isolation threshold of >50%.

#### Mathare (Starehe sub-county)

This sub-county comprises six administrative wards: Hospital, Mabatini, Huruma, Ngei, Mlango Kubwa, and Kiamaiko. The sample collection site is within the Mabatini ward, which covers 0.4 km^2^, contains eight health facilities, and had a projected population of 29,717 (15,140 male; 14,277 female) comprising 12,987 households in 2022. Mathare is among the city’s largest informal settlements, with most residents lacking access to clean piped water and formal sewer connections, resulting in wastewater discharge into the Nairobi River. The collection site is in Mathare Area 10, receiving sewage from Mathare Areas 10, 4A, and 4B, as well as from parts of the Nairobi central business district, thus representing a socioeconomically diverse catchment population.

### Sample Collection

Samples were collected using Moore swabs — autoclaved cotton gauze tied with a string and packaged in sterile ziplock bags. At each collection point, the Moore swab was inserted into the sewer and secured using the utility hole cover for a 24-hour passive collection period. Swabs were then retrieved, packaged in decontaminated ziplock bags, placed in sealed containers, and transported to the laboratory in cooler boxes with ice. Sample collection was initially conducted twice per week and subsequently increased to three times per week to achieve the target sample size of 272. This frequency was designed to account for the approximate two-day SARS-CoV-2 fecal shedding window, allowing detection of temporal variation within a given week.

### Sample Concentration and Nucleic Acid Extraction

In the laboratory, wastewater samples were concentrated using the Nanotrap® Magnetic Virus Particles (Ceres Nanosciences, USA). Nucleic acid extraction was then performed using the Qiagen QIAamp Viral RNA Mini Kit (Qiagen, Germany) following the manufacturer’s instructions. RNA extracts were immediately aliquoted: a portion was used for SARS-CoV-2 RT-PCR detection, and the remainder was stored at −80°C at the NGMSL for subsequent sequencing. RNA extraction was performed at different time points, with each batch extracted immediately or within one week of sample reception.

### SARS-CoV-2 Detection by RT-PCR

SARS-CoV-2 detection in wastewater samples was performed using the TaqPath™ COVID-19 CE-IVD RT-PCR Kit on a QuantStudio 5 Real-Time PCR System (Applied Biosystems). Samples with a cycle threshold (Ct) value below 36 were classified as positive.

### Library Preparation and Whole-Genome Sequencing

Stored RNA was thawed for library preparation using the Illumina COVIDSeq kit (10). The workflow involved: (i) cDNA synthesis using random hexamers and reverse transcriptase; (ii) amplification of cDNA using SARS-CoV-2-specific primers in two separate reactions; (iii) pooling and fragmentation of amplicons using bead-linked transposomes; (iv) adapter ligation and size selection; and (v) post-tagmentation PCR to add unique dual indexes and P5/P7 adapter sequences necessary for sequencing. Indexed libraries were pooled, purified using AMPure XP beads, quantified, normalized, denatured, and loaded onto the Illumina MiSeq™ sequencer for paired-end sequencing. Cluster generation was followed by sequencing-by-synthesis (SBS) using reversible terminator chemistry (10).

### Bioinformatic and Phylogenetic Analysis

Sequencing data were analyzed using Terra.bio and RStudio. Variant identification and frequency analysis were conducted for all sequences that passed quality filtering. A total of 181 sequences were used for variant prevalence analysis stratified by site and quarter. For phylogenetic analysis, 175 clinical sequences from GISAID (collected in Kenya between May and October 2023) and 28 wastewater sequences with ≥50% genome coverage were included. Sequences not meeting quality thresholds — including one recombinant sequence and 26 clinical sequences lacking sampling location — were excluded. Phylogenetic relationships were inferred and visualized alongside GISAID sequences from various Kenyan counties (Nairobi, Nakuru, Kiambu, and Machakos). The schematic presentation of the Laboratory workflow is presented in **figure 1**

**Figure 1:**
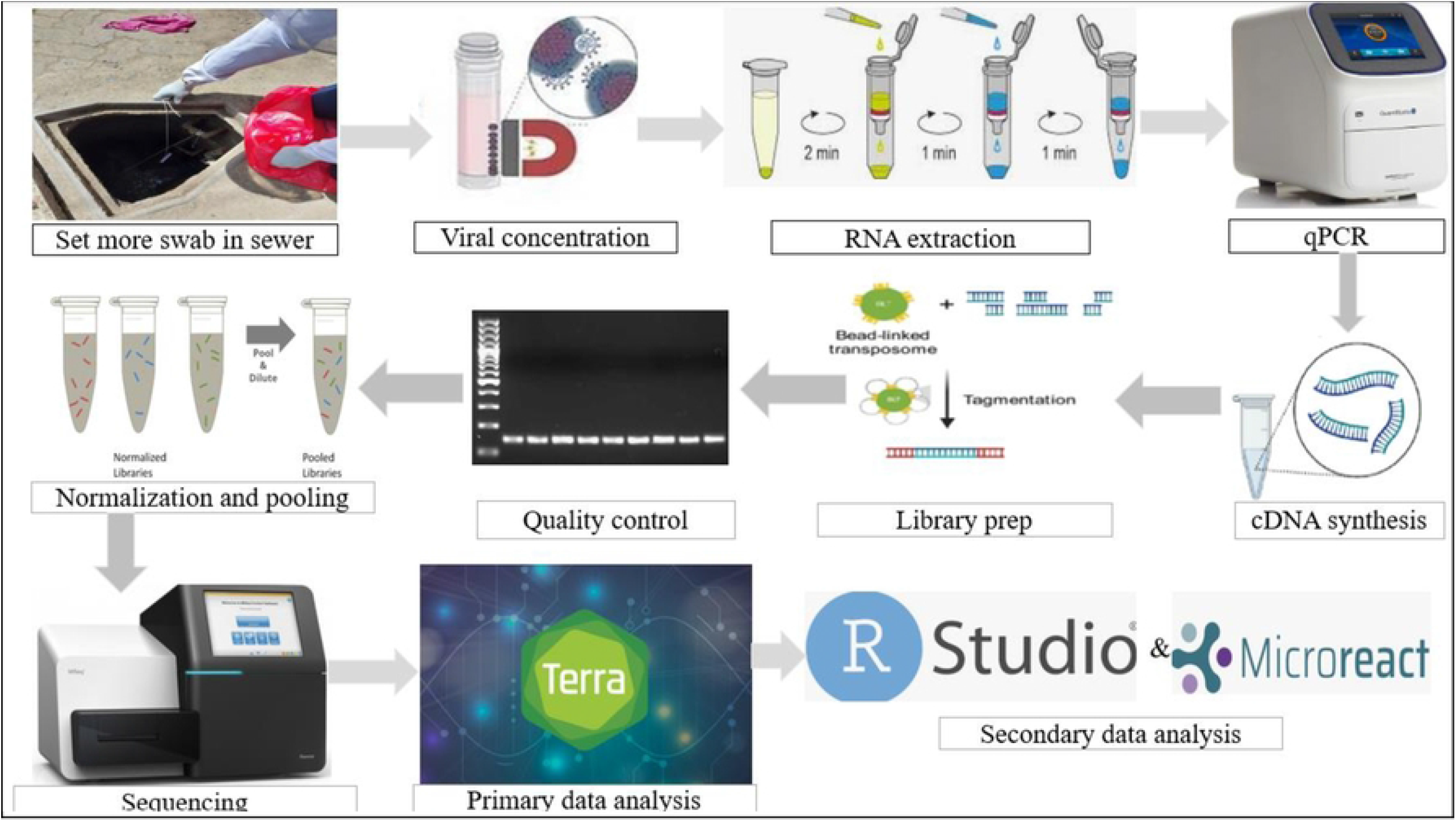
Schematic diagram of the wastewater-based genomic surveillance workflow. Overview of the end-to-end laboratory and bioinformatic pipeline employed in this study. The workflow illustrates sequential steps from field sample collection using Moore swabs, through sample concentration (Nanotrap^⨀^ Magnetic Virus Particles), nucleic acid extraction (QIAamp Viral RNA Mini Kit), SARS-CoV-2 RT-PCR detection (TaqPath^™^ COVID-19 CE-IVD RT-PCR Kit, QuantStudio 5), library preparation (Illumina COVIDSeq kit), whole-genome sequencing (Illumina MiSeq^™^), and bioinformatic analysis (Terra.bio and RStudio).

### Ethical Considerations

This study utilized wastewater samples, which are non-invasive in nature and do not contain individually identifiable information. Ethical approval was obtained from the Amref Health Africa ERC prior to study commencement. All laboratory procedures were conducted in accordance with applicable biosafety standards.

## Results

### SARS-CoV-2 Detection in Wastewater

Of the 272 wastewater samples analyzed, 238 (87.5%) tested positive for SARS-CoV-2 by RT-PCR with a Ct value of less than 36 as shown in **Figure 2**. Ct value distributions across collection dates at both sites are illustrated in **Figure 3** and it reveals the temporal variation in detection rates consistent with fluctuating community infection levels throughout the study period.

**Figure 2:**
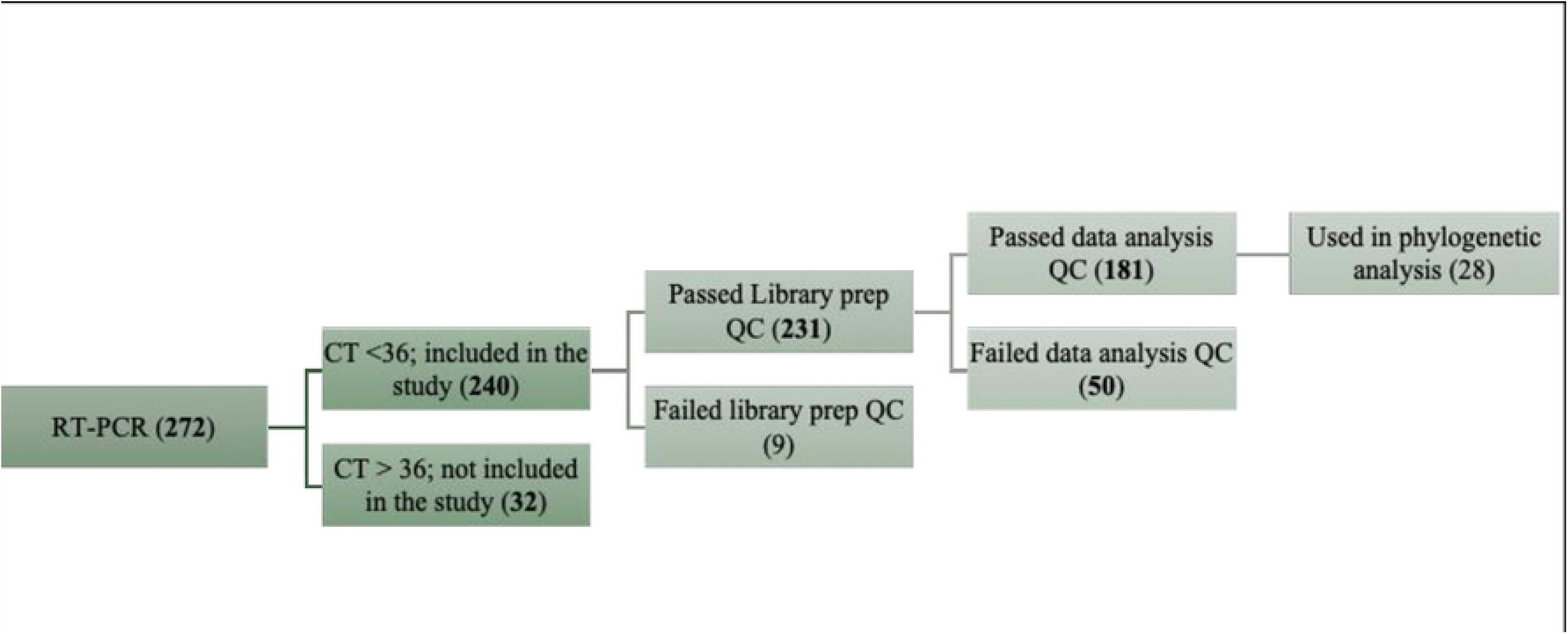
Summary of sample processing outcomes - from collection to sequencing. Flowchart depicting the disposition of all 272 wastewater samples collected during the study period. The chart shows the number of samples that were: (i) collected and processed for RT-PCR; (ii) confirmed positive (Ct < 36) or negative; (iii) subjected to library preparation; (iv) successfully sequenced; and (v) passed quality filtering for downstream variant and phylogenetic analysis (n = 181 for variant analysis; n = 28 for phylogenetics). Samples excluded at each stage are noted with reasons

**Figure 3:**
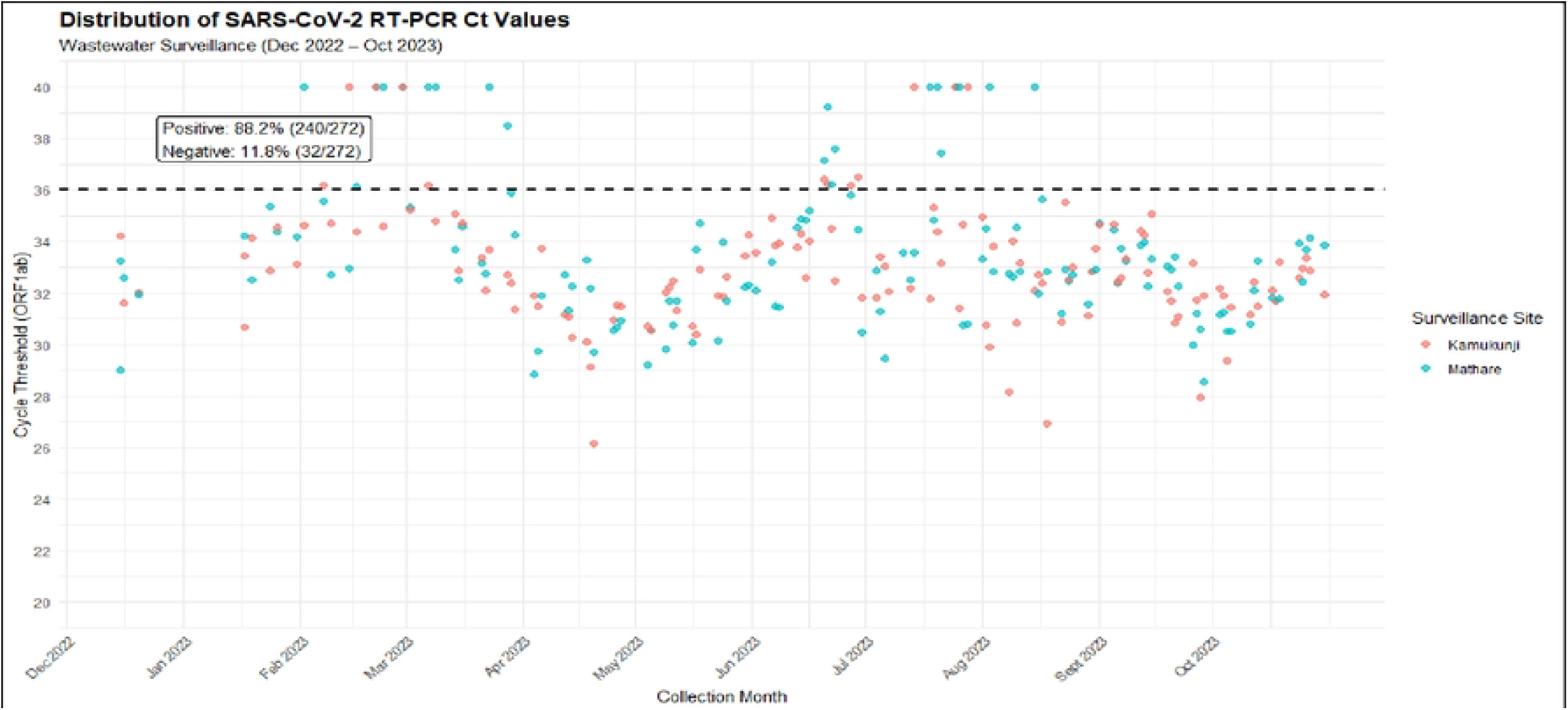
Distribution of SARS-CoV-2 RT-PCR cycle threshold (Ct) values across the study period at both surveillance sites. Time-series scatter plot displaying the Ct values of all 272 wastewater samples tested by RT-PCR from December 2022 to October 2023, stratified by collection site (Kamukunji and Mathare). The horizontal dashed line indicates the positivity threshold (Ct< 36). Temporal fluctuations in Ct values reflect variation in community-level SARS-CoV-2 infection intensity. The proportion of positive samples (238/272; 87.5%) is annotated on the figure.

### SARS-CoV-2 Variant Distribution

Genomic analysis of the wastewater sequences identified Omicron as the predominant circulating variant at both sites between January and October 2023, accounting for 59% of sequences. XBB was the second most common variant at 16%, followed by XBB.2.3 at 10%, and XBB.1.9.X at 5% **(Figure 4)**. The variant distribution observed in wastewater was concordant with that identified in 181 clinical sequences from Kenya over the same period **(Figure 5**), where Omicron (inclusive of XBB+XBB.) was also the dominant variant, XBB.2.3 was the second most diverse variant, and XBB.1.5.X, XBB.1.16.X, and XBB.1.9.X exhibited varied levels of diversity.

**Figure 4:**
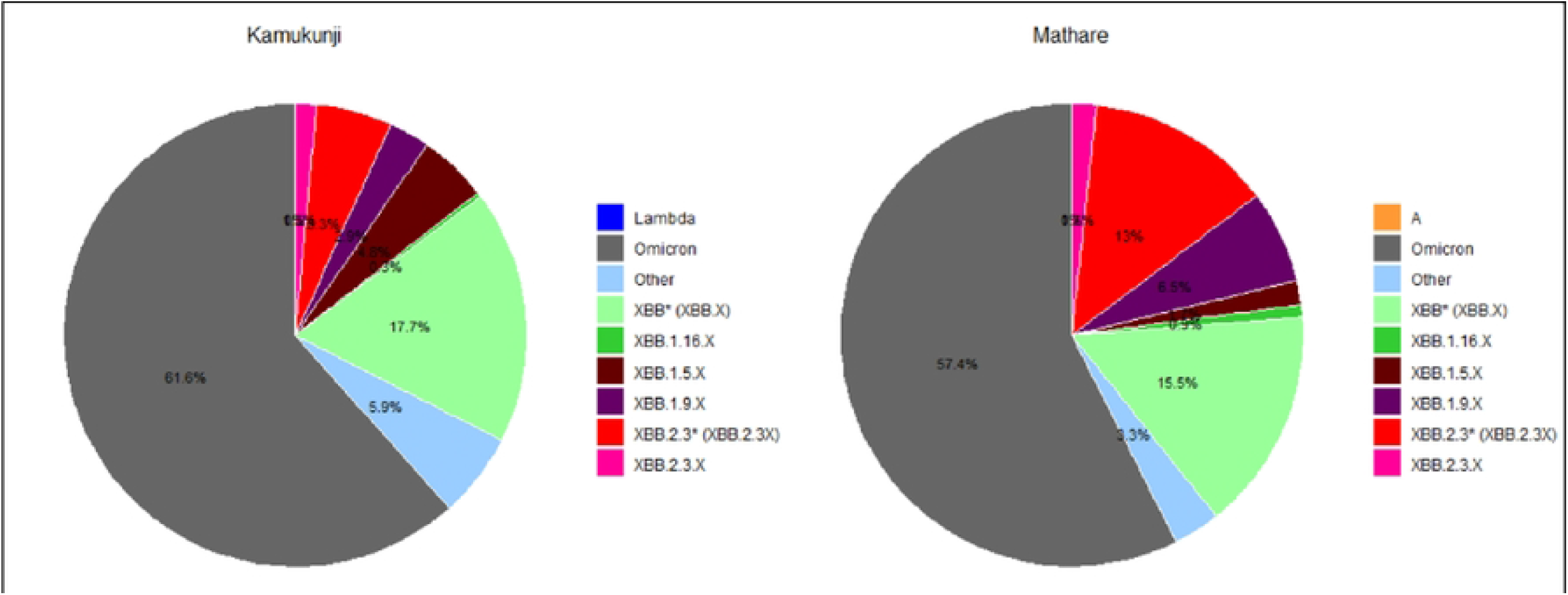
Overall distribution of SARS-CoV-2 variants identified in wastewater samples (January - October 2023). The pie chart illustrate the relative proportion of SARS-CoV-2 variants identified across all 181 genomic sequences obtained from wastewater samples at both surveillance sites combined. Variant groups include Omicron (59%), XBB.(16%), XBB.2.3 (10%), XBB.1.9.X (5%), and other minor variants. Each variant group is color-coded for visual distinction.

**Figure 5:**
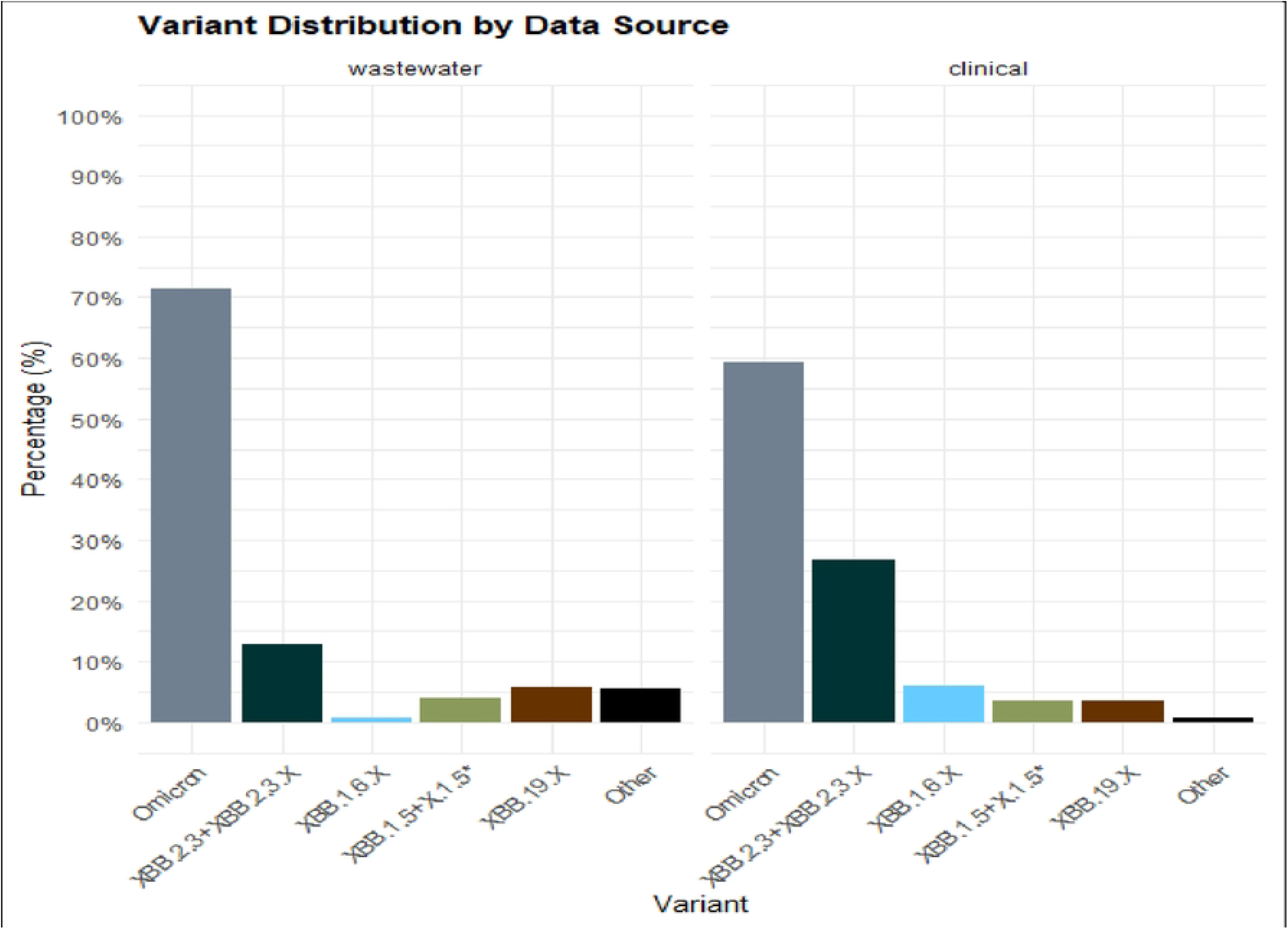
Comparison of SARS-CoV-2 variant distribution between wastewater and clinical sequences from Kenya (January-October 2023). Side-by-side stacked bar charts compare the proportional variant composition of wastewater-derived genomic sequences (n = 181) with clinical sequences available from Kenya during the same period (sourced from GISA/0). Variants depicted include Omicron (inclusive of XBB and XBB.), XBB.2.3, XBB.1.5.X, XBB.1.16.X, XBB.1.9.X, and other lineages. The figure demonstrates concordance between wastewater and clinical surveillance data

### Temporal Variant Dynamics (Quarterly Analysis)

A quarterly breakdown of variant prevalence at each site is presented in **Table 1** and **Figure 6**.

**Table 1:** Quarterly Variant Prevalence by Site (2023)

| **Table 1: Quarterly Variant Prevalence by Site (2023)** |  |  |  |  |  |
| --- | --- | --- | --- | --- | --- |
| **Variant** | **Site** | **Q1** | **Q2** | **Q3** | **Q4** |
| *Omicron* | *Kamukunji* | *0.25* | *0.28* | *0.8* | *0.6* |
|  | *Mathare* | *0.3* | *0.32* | *0.73* | *0.399* |
| *XBB.\** | *Kamukunji* | *0.18* | *0.45* | *0.08* | *0.1* |
|  | *Mathare* | *0.15* | *0.41* | *0.07* | *0.001* |
| *XBB.1.9.X* | *Kamukunji* | *0.3* | *0.05* | *0* | *0* |
|  | *Mathare* | *0.4* | *0.12* | *0* | *0* |
| *XBB.1.5.X* | *Kamukunji* | *0.2* | *0.1* | *0.002* | *0.02* |
|  | *Mathare* | *0* | *0.06* | *0* | *0* |
| *XBB.1.16.X* | *Kamukunji* | *0.02* | *0* | *0* | *0* |
|  | *Mathare* | *0* | *0.02* | *0* | *0* |
| *XBB.2.3\** | *Kamukunji* | *0.045* | *0.05* | *0.048* | *0.16* |
|  | *Mathare* | *0* | *0.02* | *0.1* | *0.5* |
| *Other* | *Kamukunji* | *0.005* | *0.07* | *0.07* | *0.12* |
|  | *Mathare* | *0.15* | *0.05* | *0.1* | *0.1* |

**Figure 6:**
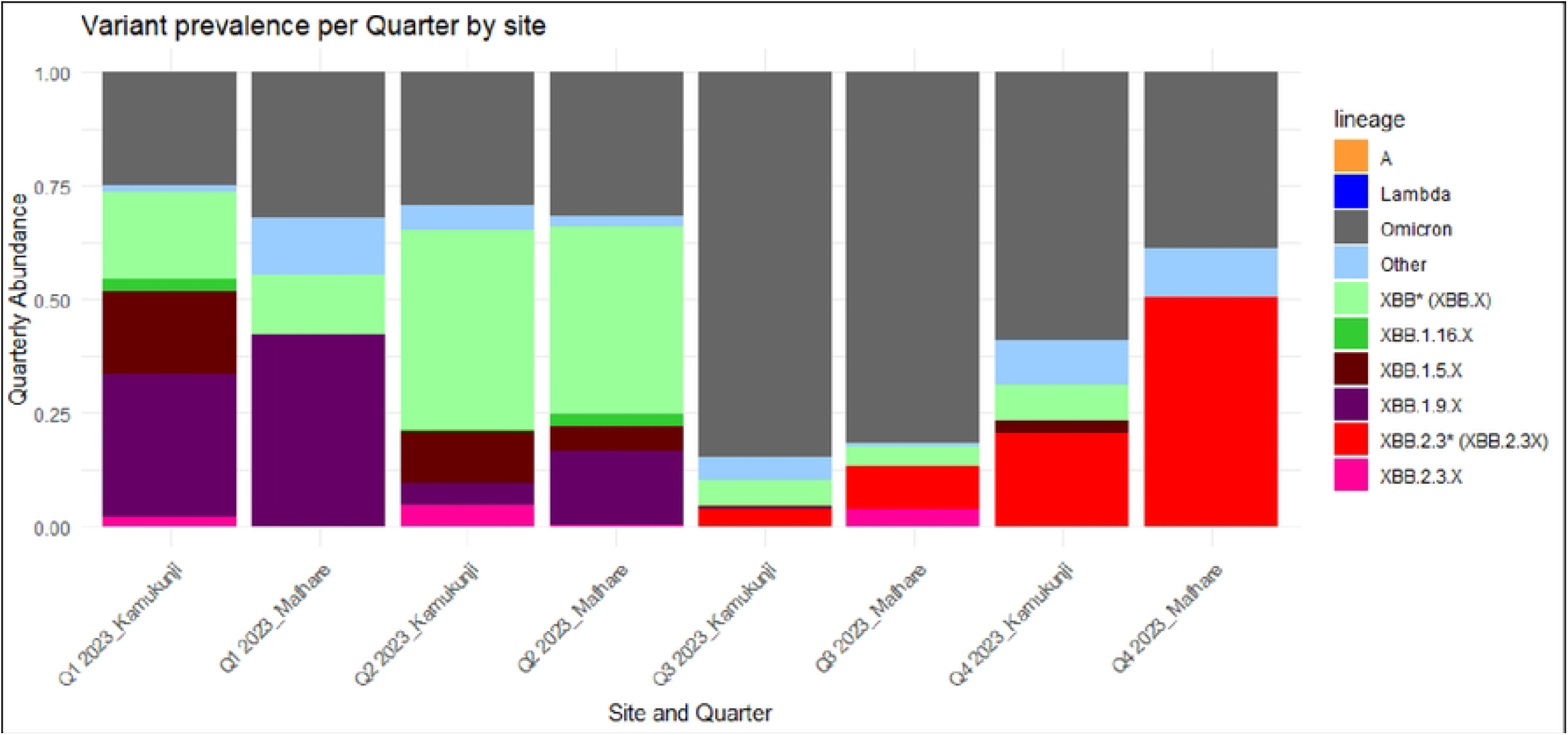
Quarterly SARS-CoV-2 variant prevalence in wastewater samples, stratified by surveillance site (2023). Grouped stacked bar charts showing the quarterly (Q1-Q4) proportional distribution of SARS-CoV-2 variants detected in wastewater at Kamukunji (Eastleigh A) and Mathare (Starehe) separately. Variants are co/or-coded consistently with Figures 6 and 7. The figure highlights the temporal emergence, peak, and decline of individual variants across study quarters and illustrates site-specific differences in variant dynamics, including the early emergence of XBB.2.3* at Kamukunji and the tater-peaking abundance at Mathare

**Figure 7:**
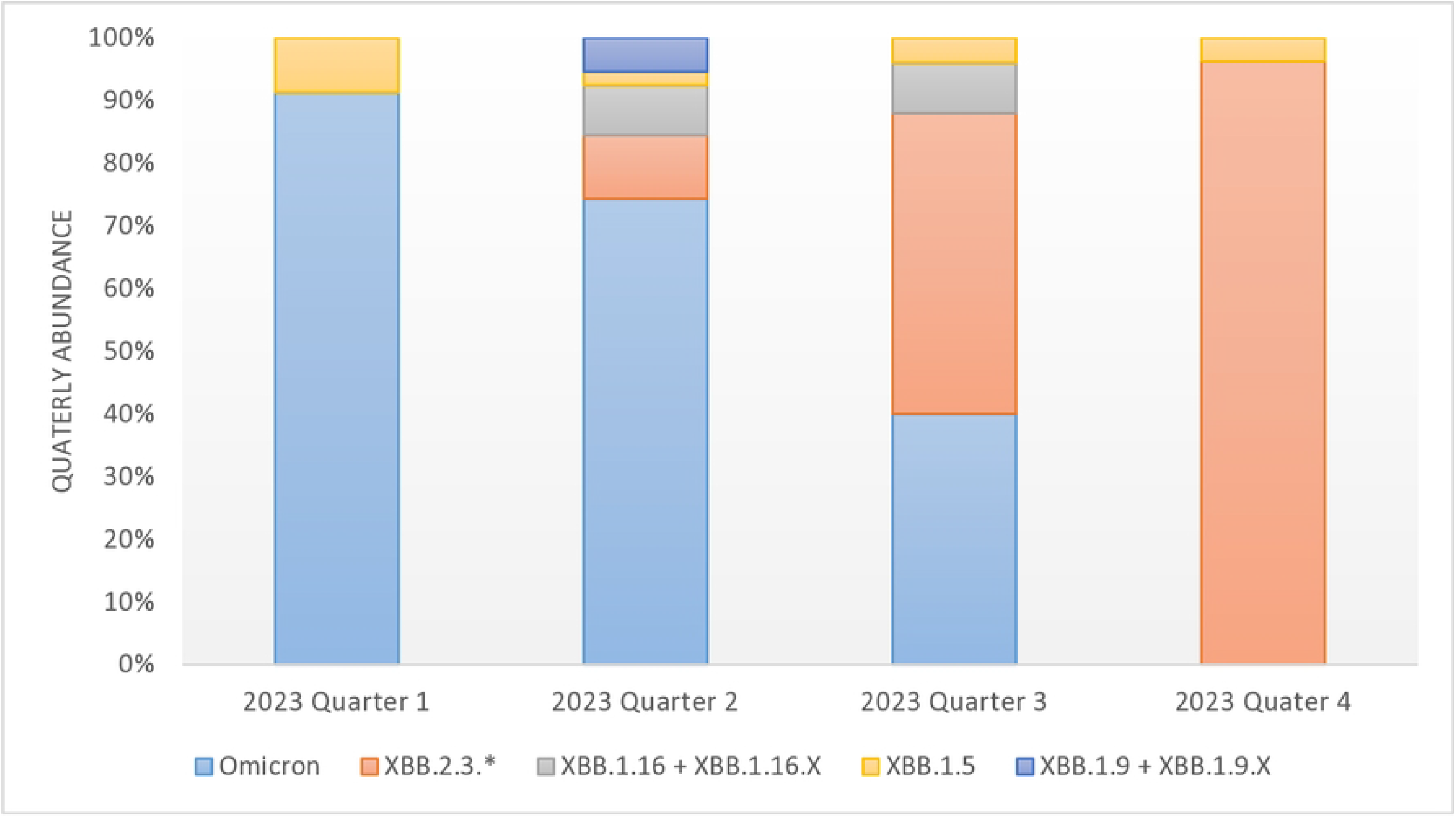
Quarterly SARS-CoV-2 variant prevalence in clinicalsequences from Kenya (2023). Stacked bar chart displaying the proportional distribution of SARS-CoV-2 variants identified in 175 Kenyan clinicalsequences obtained from GISAID, stratified by quarter (Q1-Q4, 2023). Variants depicted include Omicron, XBB.2.3*, XBB.1.5.X, XBB.1.16.X, XBB.1.9.X, and others. Quarterly clinical trends are compared with wastewater findings presented in Figure 6 to highlight concordance and instances of temporal precedence of wastewater detection

Temporal analysis of variant prevalence across the four quarters of 2023 revealed distinct circulation patterns for each identified lineage at both surveillance sites. Omicron demonstrated persistent circulation throughout all four quarters at both Kamukunji and Mathare, with its highest relative abundance recorded in the third quarter, underscoring its continued dominance within the community over the study period. XBB. followed a notably different trajectory, peaking in abundance during Q2 at both sites before declining substantially through Q3 and Q4. However, its continued detection into Q4 at Kamukunji — in contrast to its absence at Mathare during the same period — points to differential local transmission dynamics between the two communities, potentially reflecting differences in population movement, social mixing, or sewer catchment characteristics.

XBB.1.9.X was the most prevalent lineage in Q1 but exhibited a progressive decline and had disappeared entirely from wastewater at both sites by Q3. Interestingly, this variant was only identified in clinical samples during Q2, and even then, at very low abundance, suggesting that wastewater surveillance captured its earlier community circulation more comprehensively than clinical testing. XBB.2.3, by contrast, was detected at low abundance in Q1 wastewater samples from Kamukunji, while remaining undetectable at Mathare until Q2. Following its emergence at Mathare, the variant increased progressively in abundance, reaching a notably high relative proportion of 0.500 at that site by Q4. This trajectory closely mirrored trends observed in clinical sequencing data, where XBB.2.3 first appeared in Q2 and similarly increased through Q3 and Q4, reinforcing the concordance between wastewater and clinical surveillance signals. Finally, XBB.1.5.X showed a steady decline throughout the year at Kamukunji, while at Mathare it was detected exclusively in Q2, after which it was no longer identified in subsequent quarters at either site.

In clinical samples **(Figure 7**), Omicron was dominant in Q1 through Q3, with an absence in Q4 possibly attributable to insufficient clinical data during that period. XBB.2.3 first appeared in clinical samples in Q2 and increased thereafter, consistent with wastewater trends.

### Phylogenetic Analysis

Phylogenetic analysis was conducted using 175 clinical sequences from GISAID (collected in Kenya between May and October 2023) and 28 wastewater sequences with ≥50% genome coverage **(Figure 8)**. Wastewater sequences clustered within the same major phylogenetic clades as clinical sequences — 23B, 23D, 23E, and 23F — supporting the representativeness of wastewater surveillance data.

**Figure 8:**
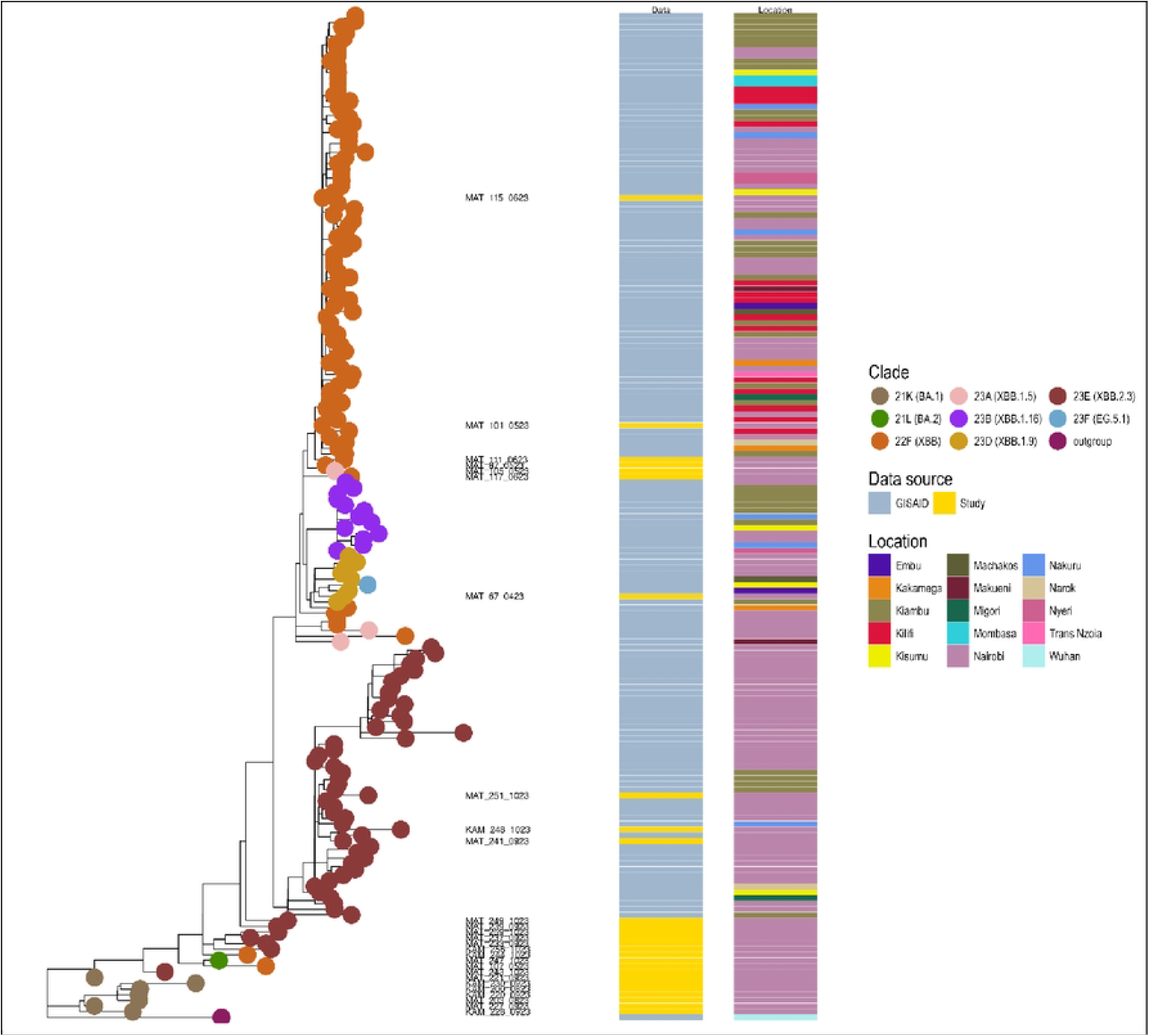
Maximum likelihood phylogenetic tree of SARS-CoV-2 sequences from wastewater and clinical surveillance in Kenya (May - October 2023). Phylogenetic tree constructed using 28 wastewater genomic sequences (≥50% genome coverage) and 175 clinical sequences from Kenya available in GISA/0, collected between May and October 2023. Wastewater sequences are color-coded and distinguished from clinical sequences. Major phylogenetic clades (238, 230, 23E, and 23F) are annotated. Sequences from Kenyan counties (Nairobi, Nakuru, Kiambu, and Machakos) are indicated by distinct symbols or colors. Notable observations - including the clustering of seven wastewater sequences within the BA.1 (21K) clade and the close clustering of XBB.2.3 wastewater sequences with contemporaneous GISAID sequences – are highlighted with annotations

Notably, seven wastewater samples collected between September and October 2023 were classified under subvariant 21K (BA.1), placing them earlier in the phylogenetic tree without clustering closely to contemporary clinical sequences. This suggests that this lineage remained in circulation within the community as late as October 2023, a finding not captured in available clinical data — demonstrating the capacity of wastewater surveillance to detect variants that may be missed by clinical testing. Other wastewater sequences collected between September and October 2023 were classified as XBB.2.3 and XBB. and clustered closely with GISAID sequences from Nairobi, Nakuru, Kiambu, and Machakos, affirming that our sequences represent circulating variants in Kenya. Samples collected between April and June 2023 were classified as XBB. and clustered alongside GISAID sequences from across Kenya.

## Discussion

### Principal Findings

This study demonstrates the feasibility and utility of wastewater-based genomic surveillance for tracking SARS-CoV-2 variant dynamics in two densely populated informal urban settlements in Nairobi, Kenya. Between January and October 2023, Omicron was the dominant variant detected in wastewater samples from both Kamukunji and Mathare, accounting for 59% of sequences — a finding consistent with clinical surveillance data from Kenya over the same period (11) (12).

The predominance of Omicron is further supported by Musundi et.al (2024), who reported FY.4 (XBB.1.22.1), a sub-lineage classified under Omicron in Kenyan clinical and wastewater sequences, as the dominant circulating variant in Kenya between April and July 2023(13). This corroborates our finding that Omicron and its subvariants constituted the major circulating lineages across all regions of Kenya in 2023.

### Wastewater as an Early Warning System

A critical observation in this study was the detection of XBB.2.3 in wastewater samples from Kamukunji in Q1 at low abundance — a period during which this variant was absent in clinical samples. This variant subsequently appeared in Mathare wastewater in Q2, and was not detected in clinical samples until Q2. This temporal precedence of wastewater detection over clinical detection illustrates the early warning capacity of WBE, allowing public health authorities to anticipate and prepare for emerging variant circulation before it manifests in clinical case counts (6) (7). Similarly, the detection of BA.1 (21K) in wastewater samples as late as September–October 2023, in the absence of corresponding clinical detections, suggests that wastewater surveillance captures a more comprehensive picture of circulating lineages — including those underrepresented or absent from clinical testing — than clinical surveillance alone (8).

### Site-Specific Variant Dynamics

Comparative analysis between Kamukunji and Mathare revealed some site-specific differences in variant timing and abundance. For instance, XBB. persisted into Q4 at Kamukunji but disappeared from Mathare by Q3. XBB.2.3 was detected earlier in Kamukunji and later reached a notably higher abundance in Mathare in Q4 (0.500 vs. 0.160). These differences may reflect distinct population mixing patterns, mobility dynamics, or healthcare-seeking behaviors across the two communities. They may also be influenced by the broader geographic catchment of the Mathare site, which receives wastewater from multiple socioeconomically diverse zones, including parts of the Nairobi central business district (12).

### Concordance with Clinical Surveillance Data

Phylogenetic analysis confirmed that wastewater sequences clustered within the same major clades (23B, 23D, 23E, 23F) as clinical sequences from Kenya. Furthermore, XBB.2.3 was identified as a dominant circulating variant in Kenya in 2023, as documented by Hodcroft et al. (2024)(14), consistent with its progressive increase in both wastewater and clinical samples. The alignment of our sequences with GISAID-representative sequences from Nairobi, Nakuru, Kiambu, and Machakos further validates the representativeness of our wastewater genomic data.

### Strengths

The key strength of this study lies in the demonstration that wastewater surveillance can accurately reflect variant diversity within a defined community, including variants not captured by clinical testing. The use of WHO-validated surveillance sites, rigorous laboratory protocols, and triangulation of findings with independent GISAID clinical data strengthens the credibility and generalizability of the findings. The study’s prospective design and consistent sampling schedule further support the reliability of temporal trend analyses.

### Limitations

Several limitations must be acknowledged. First, the RNA extracts were subjected to extended storage (up to one year at −80°C) before sequencing, which resulted in RNA fragmentation and reduced genome coverage during data analysis. It is recommended that wastewater-extracted nucleic acids be sequenced within six months of storage at −80°C to minimize degradation. Second, the absence of viral quantification (only Ct values were used to define positivity) precluded an analysis of the relationship between viral load and sequencing success rates. Future studies should incorporate quantitative PCR to enable viral load estimation. Third, clinical data could not be obtained from the specific catchment areas served by the two surveillance sites, limiting individual-level correlation between wastewater and clinical findings. Such paired data would significantly enhance the inferential value of comparative analyses. Fourth, the study cannot be generalized to represent Kenya as a country because it relied only on two environmental surveillance sites in one city, which limits extrapolation to the broader Kenyan or African context.

## Conclusions

This study provides evidence that wastewater-based genomic surveillance is a viable, sensitive, and population-representative tool for tracking SARS-CoV-2 variant dynamics in resource-limited urban settings. The detection of Omicron as the predominant circulating variant and the concordance of wastewater findings with clinical sequencing data from Kenya demonstrate the reliability of this approach. Critically, wastewater surveillance provided earlier signals of variant emergence than clinical data, underscoring its value as an early warning system for public health response.

Given the non-invasive nature, cost-effectiveness, and broad population coverage of wastewater-based epidemiology, several recommendations are put forward to strengthen and expand its application in Kenya and beyond. Wastewater-based surveillance should be formally scaled up across multiple sites to generate nationally representative data, with future studies incorporating quantitative SARS-CoV-2 measurement alongside genomic analysis to enhance the depth of surveillance findings. The scope of WBE should further be broadened beyond SARS-CoV-2 to include other pathogens of public health significance, such as Influenza A and B, Adenovirus, Vibrio cholerae, Measles, and Salmonella typhi, while also serving as a community-level tool for monitoring antimicrobial resistance (AMR) gene signatures. From a methodological standpoint, nucleic acid sequencing should be performed within six months of RNA extraction to preserve genomic integrity and maximize genome coverage in subsequent analyses.

## Data Availability

All data produced in the present work are contained in the manuscript

